# Can vocational advice be delivered in primary care? The Work And Vocational advicE (WAVE) mixed method single arm feasibility study (NCT04543097)

**DOI:** 10.1101/2025.09.15.25335634

**Authors:** G Wynne-Jones, G Sowden, I Madan, K Walker-Bone, CA Chew-Graham, B Saunders, M Lewis, K Bromley, S Jowett, V Parsons, G Mansell, K Cooke, SA Lawton, C Linaker, J Pemberton, C Cooper, NE Foster

## Abstract

**Objectives:** Most patients with health conditions necessitating time off work consult in primary care. Offering vocational advice (VA) early within this setting may help them to return-to-work (RTW) and reduce sickness absence. Previous research shows the benefits of VA interventions for musculoskeletal pain in primary care, but an intervention for a much broader primary care patient population has yet to be tested. The WAVE feasibility study tested patient identification and recruitment methods, explored participants’ experiences of being invited to the study and their experiences of receiving VA.

**Design:** A mixed method, single arm feasibility study comprising both quantitative and qualitative analysis of recruitment and participation in the study.

**Setting:** Primary care

**Methods:** The study included participant follow-up by fortnightly SMS text and 6-week questionnaire. Stop/go criteria focused on recruitment and intervention engagement. The semi-structured interviews explored participants experiences of recruitment and receipt and engagement with the intervention.

**Results:** Nineteen participants were recruited (4.3% response rate). Identification of participants via retrospective fit-note searches was reasonably successful (13/19 (68%) identified), recruitment stop/go criteria were met with ≥50% of those eligible and expressing an interest recruited. The stop/go criterion for intervention engagement was met with 16/19 (86%) participants having at least one contact with a Vocational Support Worker (VSW). Five participants were interviewed; they reported positive experiences of recruitment and felt the VA intervention was acceptable.

**Conclusion:** This study demonstrates that delivering VA in primary care is feasible and acceptable. To ensure a future trial is feasible, recruitment strategies and data collection methods require additional refinement.

***Trial registration:*** Clinical Trials: NCT04543097

***Protocol number:*** Version 5.1

**Article summary:**

- This is the first study to test the feasibility of delivering a VA intervention to patients who present in primary care, regardless of their health condition.
- The study used mixed methods to fully explore feasibility of the delivery of a full trial
- The findings can usefully inform the development of the methods for a future trial to ensure that it meets the needs of participants in supporting them to return-to-work after a period of absence

## Background

Absence from work is increasing across European counties each of which has their own models for managing this absence (1). The availability and provision of vocational advice in the UK is variable and often only accessible to those working for larger organisations (2). It is estimated that just 45% of all employees in the UK have access to occupational health, which is lower than comparable countries (3). The Office for National Statistics (ONS) reported that 2.5 million people are reporting long-term sickness as the reason for not being in work; this has risen by 500,000 since 2019 and is not thought to be solely related to the COVID-19 pandemic (4). Of concern, long-term sickness absence is rising fastest in younger age groups (aged 25-34 years) meaning that individuals potentially have an extended or even permanent period of economic inactivity due to sickness. Furthermore, the ONS reported that just 16% of those on long-term sickness absence returned to employment between 2021-2022 (4). Analyses of new claimants applying for the UK unemployment benefit for sick and disabled individuals (Employment and Support Allowance) found that 61% of claimants had sickness absence from their last job and 75% had decided to stop working altogether (5). Good quality, timely, vocational advice and support in primary care may improve these outcomes (6), improve patients’ health and quality of life and benefit wider society by active engagement in the workforce (7). Whilst general practitioners (GPs) have been expected to have an active role in advising and supporting patients back into work, a role for other health professionals and non-health professionals in managing this interface is recommended (8,9). Reforms to UK legislation introduced in 2022 now authorises other clinicians (nurses, physiotherapists, occupational therapists, and pharmacists) to issue and certify fit notes (previously referred to as the sick note) (10). However, fit notes are often issued via an online request and many patients receive a fit note without speaking to a clinician about their health and work (11).

It is important that people are directed to timely, evidence-based sources of support and advice about their health and work issues (e.g., to address obstacles to RTW) before they get to the stage where they become long-term absent from the workforce. The UK Government’s report “Health is everyone’s business” highlights the need for improved vocational advice and support to be offered as part of economic recovery plans (12). The report focuses on the need to develop and deliver new occupational health models and make greater use of technology to support small and medium enterprises to access occupational health services. The WAVE study aimed to address these challenges, by determining the feasibility of a new model of delivery of vocational advice (to support occupational health) via telephone and videoconference, addressing the need for a flexible and technology-based delivery.

This feasibility study is reported in line with the CONSORT extension for feasibility and pilot trials (13).

## Objectives

i. To test patient identification methods, approach to screening for eligibility and recruitment, and people’s willingness to engage with a Vocational Advice (VA) intervention in a single group feasibility study
ii. To test data collection processes, for response rates and completeness of data
iii. To understand participants’ experiences of being invited to the study, the delivery of the VA intervention, and the usefulness of the intervention in supporting them to RTW through semi-structured interviews reported in this paper and consultation recordings (published elsewhere (14))

## Methods

The protocol for the feasibility study is published in full on the National Institute for Health and Care Research (NIHR) journals library (15). Patients and the public were involved in the development of the research question and at all stages of the design and delivery of the feasibility study.

## Design

A mixed method, single arm feasibility study comprising both quantitative and qualitative analysis. With, stop/go criteria to assist decision making about whether to proceed to a full trial, with linked semi-structured interviews.

## Setting

The study screened and recruited participants in primary care with recruitment taking place in general practices in three regions in England: Staffordshire, Wessex and South London.

## Inclusion and exclusion criteria

Eligibility criteria were informed by subgroup analyses of the previous Study of Work And Pain (SWAP) trial data which suggested that the VA intervention may be more effective in those participants who had at least two weeks absence from work (16).

### Inclusion criteria

1. adults aged 18 years and over
2. currently in paid employment (full or part time)
3. currently absent from work for at least two consecutive weeks but not more than six continuous months
4. receiving a fit note
5. have access to a mobile phone that can receive and respond to SMS text messages
6. able to read and write English
7. able to give full informed consent
8. willing to participate.

### Exclusion criteria

1. long-term work absence defined as longer than six continuous months
2. pregnant or on maternity leave
3. patients presenting with signs or symptoms indicative of serious illness requiring urgent medical attention (‘red’ flags)
4. severe mental health problems (e.g. severe depression with risk of self-harm, exacerbation of schizophrenia or bipolar disorder, cognitive impairment or lack of capacity) high vulnerability (e.g. palliative stages of illness, recent bereavement, dementia)

## Identification and recruitment of potential participants

To test patient identification methods, approach and eligibility for the WAVE study, several recruitment methods were assessed. Recruitment took place between December 2021 and March 2022. A detailed description of the methods of identification and recruitment are reported in the protocol and are summarised below (17).

### Method A: Identification through automated health informatics IT Protocol during “real time consultations”

Potentially eligible patients were identified using an automated medical record protocol (a “pop-up”) which activated when the clinician completed an electronic fit note (eMED3) *during* a consultation with a patient. This pop-up only triggered if the patient met the inclusion/exclusion criteria. The clinician would then ask the patient for consent to share the patient contact details with Keele Clinical Trials Unit (CTU), who then posted the patient a study pack inviting them to the study.

### Method B: Identification through searches of the general practice medical record after consultation where a pop-up is used to assess eligibility on completion of a fit note

Potentially eligible patients were identified using a pop-up which activated when the clinician completed an electronic fit note (eMED3) *after* a consultation with a patient. This pop-up only triggered if the patient met the inclusion/exclusion criteria. Patients identified as being potentially eligible were then sent a study pack from the practice to invite them to the study.

### Method C: Identification through retrospective searches of the general practice medical record for all fit notes

The final method was designed to reduce the interruption to consultations that pop-ups bring. Clinicians issued fit notes as usual and a search of patients who had been issued a fit note within the past 7 days was undertaken, with clinicians screening the lists identified to assess eligibility. Practices then sent a study pack to patients inviting them to the study.

For all recruitment methods, on receipt of the study pack, potential participants were asked whether they were still absent from work because of their health condition. Those that were still absent were asked to complete the consent form and baseline questionnaire and return them to Keele CTU.

## Data collection

To test patient identification methods and approach to screening for eligibility and recruitment the number and proportion of potentially eligible patients identified, invited and consenting to participate was collected via an audit of each of the recruitment methods. Willingness to engage with the VA intervention, the take up of the offer of the intervention, and the steps of the intervention subsequently engaged with were identified from the case report forms which described the intervention delivery for each participant.

Self-reported data were collected via postal questionnaire at baseline and at six-weeks follow-up and by SMS messaging every two weeks for a period of six-weeks. This data collection allowed the processes to be assessed, and any data collection issues related to completeness of data, to be identified and adapted should the study progress to a full trial. The following data were collected:

### Work absence

Participants were asked to report the total number of days absence due to their health condition in the preceding six-weeks alongside their current work status. RTW was collected by contacting participants on a fortnightly basis by SMS. Contact was maintained until a sustained RTW was achieved (defined as return to *any work* for at least four weeks). RTW was measured via SMS text message using the following questions:

⍰ On a scale of 0 to 10, where 0 is very poor and 10 is very good, how would you rate your general health over the past 2 weeks?
⍰ Have you returned to work? Yes/No
⍰ If yes, on which date did you return to work e.g. 13SEP2021?

Work interference was measured using the Work Productivity Activity Impairment (WPAI) questionnaire (18). Work performance was measured using the Single Item Performance Question (SIPQ) (19).

### Additional measures

A number of measures corresponding to concepts considered important and included in the logic model underpinning the VA intervention were also measured, the publication describing the development of the logic model is in progress. These are reported in full in the protocol but included (17):

### Personal health

Physical health and mental health were measured using the Short Form 12 (SF12) (20,21). Depression was measured using the Patient Health Questionnaire (PHQ) 8 (22), anxiety was measured using the Generalised Anxiety Disorder (GAD) 7 (23) and quality of life was measured using the EQ-5D-5L (24).

### Personal influences

The attitudes and beliefs about work questionnaire, used in a previous randomised controlled trial (RCT), was developed to assess how participants view working with health conditions (16). The Return-to-Work Self-Efficacy (RTW-SE) (25) questionnaire measured changes in a participant’s confidence to return to work.

### Personal behaviours

Physical activity level was measured using the General Practice Physical Activity Questionnaire (GPPAQ) (26).

### Occupational measures

Work absence in the past 12 months, use of other work support services (provided through health services or the participants workplace), current job title and characteristics, perceived global stress at work (27), satisfaction with work (28) and how soon the participants expected to resume their normal job without any limitations.

Lastly, participants reported the main health condition (mental ill-health (MH), musculoskeletal conditions (MSK) or other condition) that resulted in their work absence, and sociodemographic data.

## Engagement with the VA intervention – linked semi-structured interviews

To understand participants’ experiences of being invited to the study, the delivery of the VA intervention and the usefulness of the intervention in supporting RTW, all participants recruited to the feasibility study were invited to participate in a semi-structured interview. On receipt of a completed reply slip the study team contacted the participant by phone to arrange a suitable date and location for the interview. A consent form was completed prior to each interview, either written if face-to-face or audio-recorded consent where interviews were undertaken by phone or video. Topic guides were used to support the interviews and included questions about the individual’s work absence, the acceptability of participant information about the WAVE study and the VA intervention, their experience and views of the recruitment process and their experiences of the VA intervention delivery, content and usefulness.

Engagement with the VA intervention was further assessed through audio recordings of consultations between participants and VSWs. These data informed the feasibility study and are reported elsewhere (14).

## Description of the intervention

The intervention delivered was a work-focused VA intervention remotely delivered by vocational support workers (VSWs). The development of the intervention and the training package for VSWs forms the focus of a separate publication currently in progress. In summary, the intervention was based on a logic model detailing key treatment targets (obstacles to RTW) including personal factors: health; cognitions; behaviors; emotions, and occupational factors including workplace contact; communication; and workplace adjustments. Intervention processes focused on supporting participants to tackle obstacles to RTW and included methods such as goal setting; problem solving; case management; and RTW planning. The intervention was delivered using the principles of stepped care and case management with the VSWs stepping up the intervention when necessary and taking on a case manager role to support participants’ RTW.

## Sample size

No formal sample size was calculated for the feasibility study; however, the aim was to recruit up to 30 participants, with approximately 10 from each of the three geographical regions. For the semi-structured interviews these participants were all invited to an interview.

## Analysis

To assist decision making about whether to proceed to a full trial, the main stop/go criteria in the feasibility phase were:

i. Recruitment uptake: uptake of those eligible and who expressed an interest in the study: <25% (red); 25%-49% (amber); and ≥50% (green)
ii. Engagement with the VA intervention: the percentage of patients who had at least one contact with a VSW <40% (red), 40%-65% (amber), and >65% (green).

Descriptive analysis of the key feasibility measures were:

- Number (percent) of patients who were identified in each identification method
- Number (percent) of patients who were eligible and interested in taking part in the study
- Number (percent) of patients who consented to participate
- Number (percent) of study participants who engaged with each step of the study intervention
- Completeness of questionnaires at baseline and six-months and completeness of SMS text messaging responses

### Analysis of interviews

The audio-recordings of interviews were transcribed in full and anonymised through replacing names with pseudonyms and removing other potentially identifiable information. Data were analysed through an inductive, exploratory framework using thematic analysis and informed by the constant comparison method, looking for connections within and across interviews, and across codes, highlighting data consistencies and variation (29,30). Whilst it was intended that data collection and analysis be driven by saturation, defined as ‘informational redundancy’ (31), the final sample size was restricted to the small number of participants in the feasibility study who returned a reply slip in response to the interview invitation.

Anonymised transcripts were systematically coded on a line-by-line basis by the same qualitative researcher who conducted the interviews (BS), with the aid of the software program Nvivo 10, to identify recurrent concepts inductively. Coding was reflexive and recursive, with codes being revisited considering the findings of subsequent data collection. Three interview transcripts were independently coded by another member of the research team (CCG). Both coders have significant experience in qualitative analysis and brought different disciplinary perspectives to the data (BS medical sociology; CCG academic general practice). The aim of independent coding was therefore to understand cross-disciplinary perspectives on the data and, through discussion, to come to an agreement on shared meanings and interpretations.

## Results

### Feasibility criterion

The flow of patients through the feasibility study is shown in Figure 1. Across the three methods of patient identification, 445 patients were screened as potentially eligible. Method C (identification through retrospective searches of fit notes in the medical record) was the most successful with 366 (82%) patients identified and mailed a study pack, compared to 53 (12%) in Method A (identification through automated searches during real time consultations) and 26 (6%) in Method B (identification through searches of the medical record after a consultation where a pop-up is used to assess eligibility).

**Figure 1:**
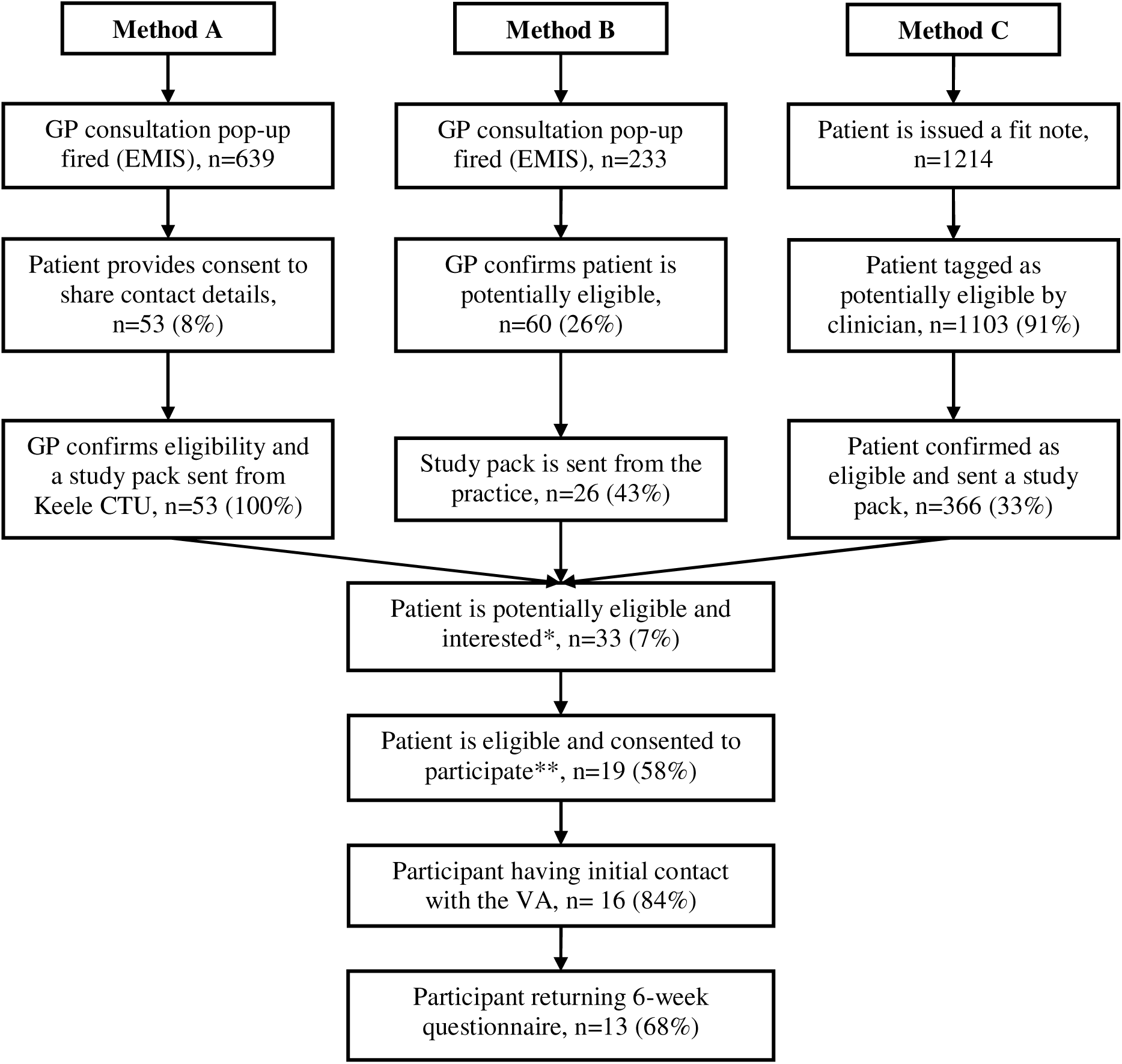
Flow diagram showing the flow of patients throughout the study * The patient has returned their study pack containing their consent form and/or questionnaire. *Potentially eligible refers to the patient being determined as potentially eligible to take part in the study according to the site criteria (i.e., fulfilling all inclusion/exclusion criteria at study site assessment). Interested is defined as returned full/partial completion of either or both the baseline questionnaire and/or consent form*. ** The patient has confirmed in their questionnaire that they are eligible as they have not yet returned to work (‘yes’ response to question A1 confirming that the patient is still absent from work) and provided full consent to participate in the study.

Of those mailed a study pack the number of participants potentially eligible and interested measured through return of either or both their consent form and baseline questionnaire was low at 33 (7%) across all recruitment methods. Of these, 19 participants (58%) were screened as absent from work, consented and recruited to the feasibility study. This was a green signal (i.e. >50%) (by progression criteria (i)). However, this gave an overall recruitment response rate of 4.3%.

The second feasibility criterion focused on engagement with the VA intervention measured through the number (%) of study participants who engaged with each step of the WAVE vocational advice intervention. Case report forms, documenting intervention delivery, were available for 14 participants. Of these 14, 12 participants (86%) engaged with the intervention successfully, with 4 (33%) reaching step 1, and 8 (76%) reaching step 2 of the intervention, none of the participants reached step 3 (Table 1).

**Table 1:**
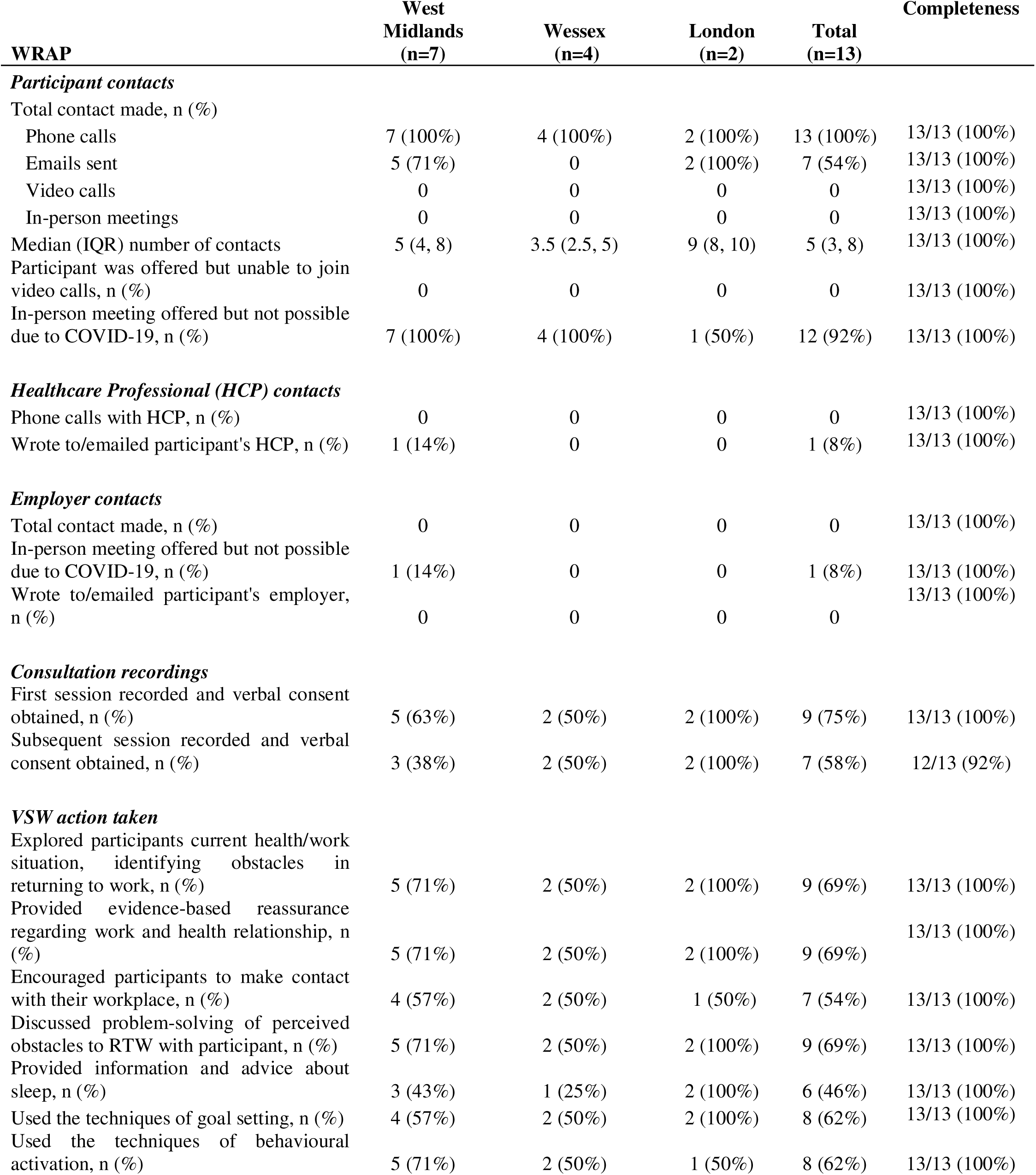
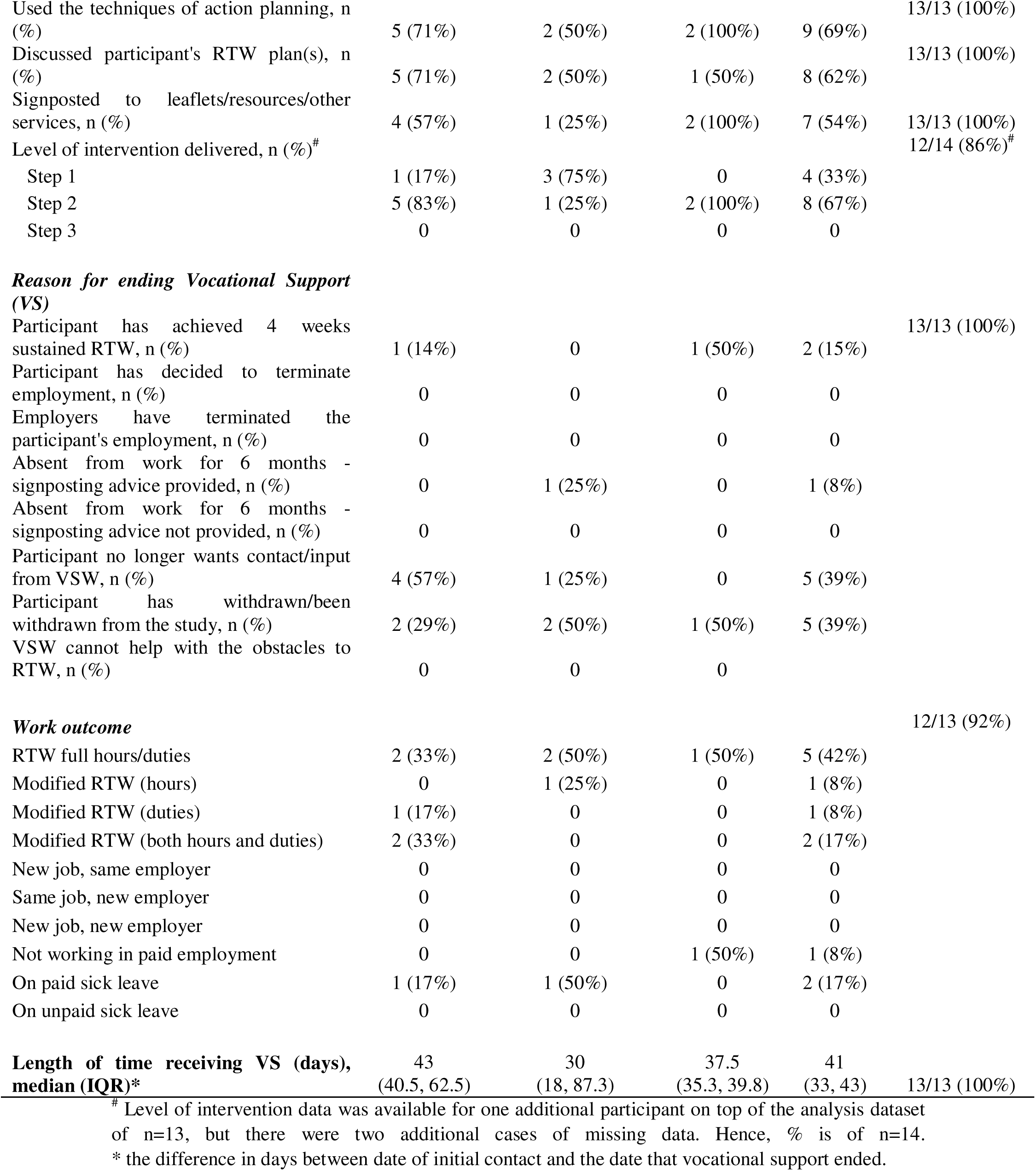
Summary of information recorded in the WAVE Return to work Assessment and action Plan (WRAP) Case Report Form

To test feasibility of processes the response rate at six months and completeness of data in the questionnaires at baseline and six months and the SMS text message data were evaluated.

Overall, there were high levels of completeness for the baseline questionnaire with the exception of two items on the WPAI questionnaire (18) where participants struggled to complete the questions around presenteeism and work productivity (Table 2 reports a summary of baseline data with Supplementary Table 1 reporting the full dataset). At six-weeks follow-up 13/19 (68%) participants returned their questionnaire (Figure 1). Again, completion of the questionnaire was high with all participants completing questions on measures of absence and work status that are likely to be primary outcomes in a full trial (Table 3 reports a summary of six-week data with Supplementary Table 2 reporting the full dataset). Furthermore, the completion of the performance and presenteeism questions of the WPAI (18) had improved.

**Table 2:**
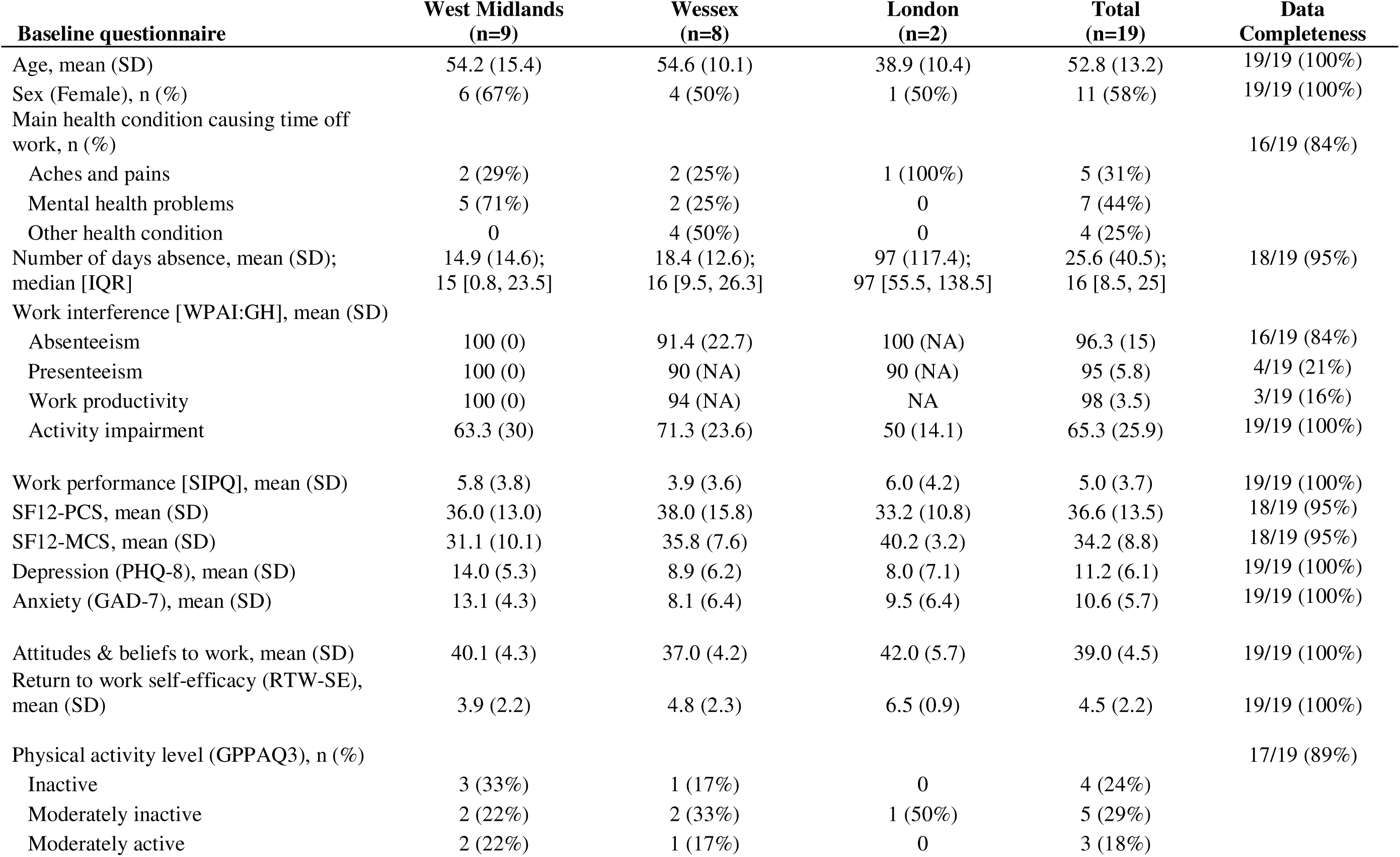
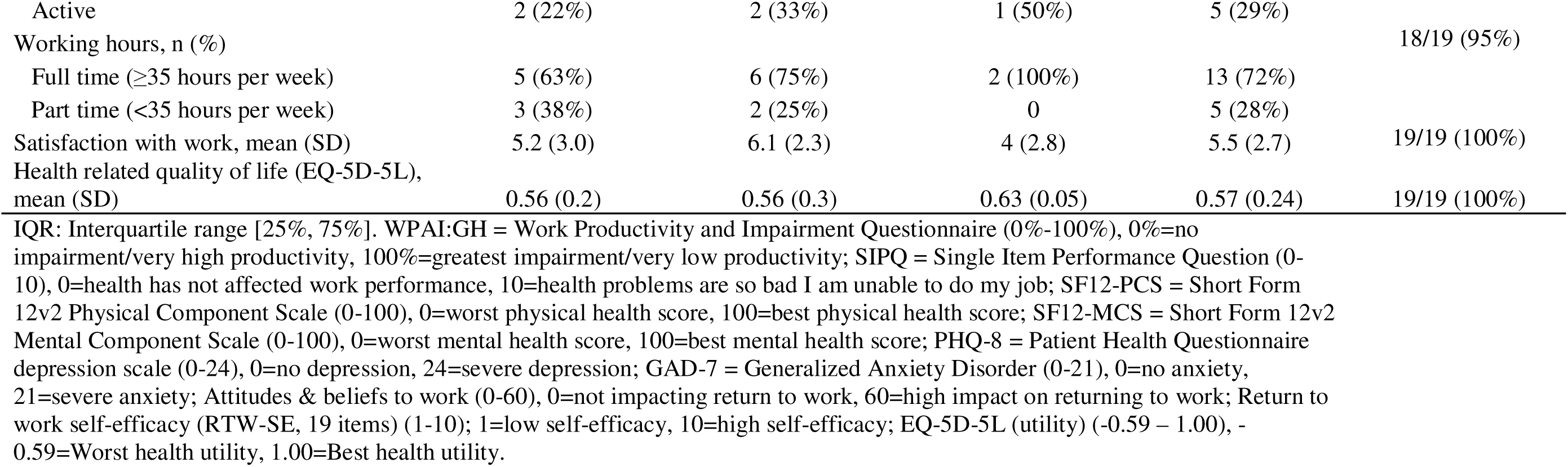
Summary of participant characteristics at baseline

**Table 3:**
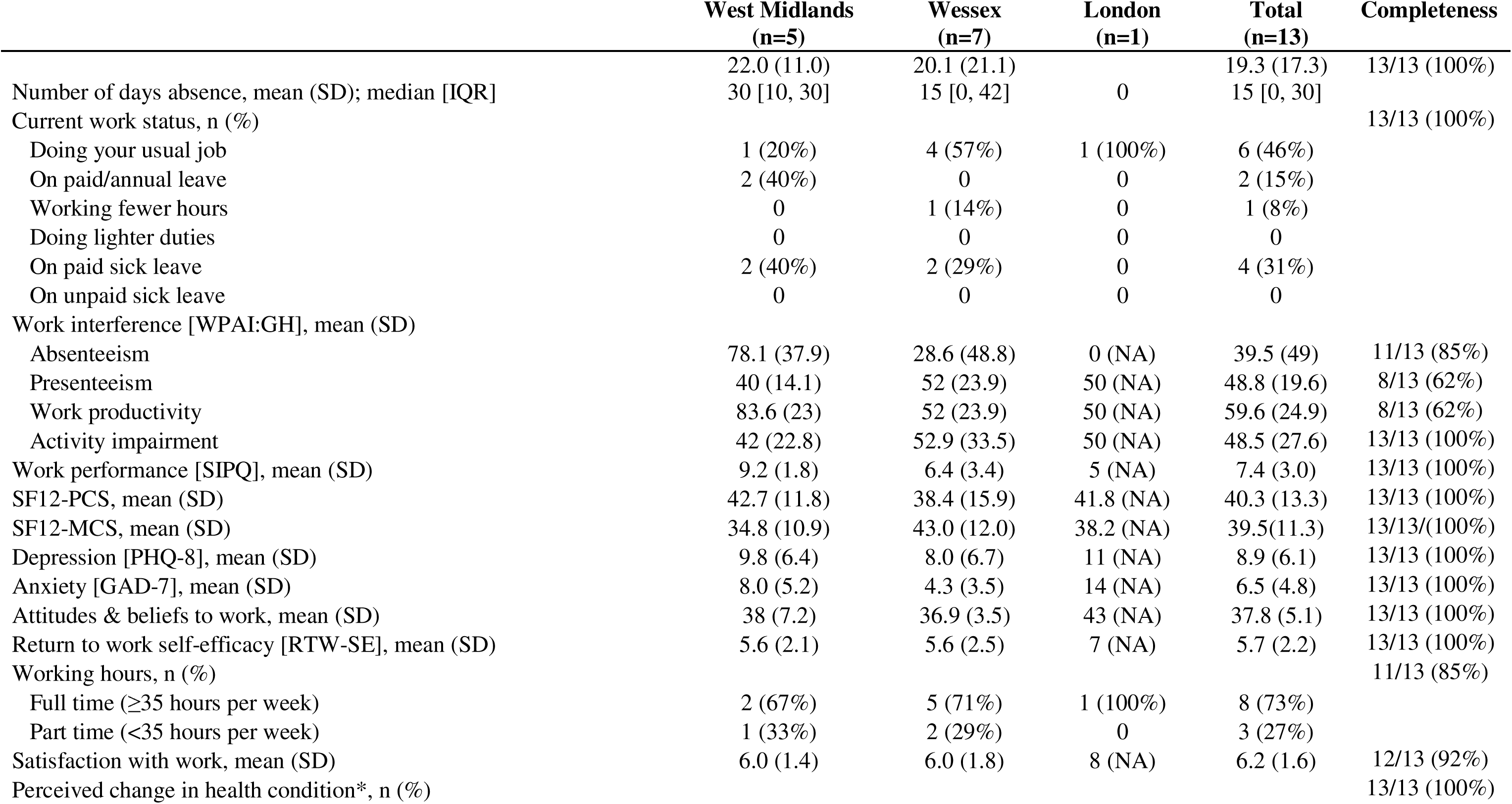
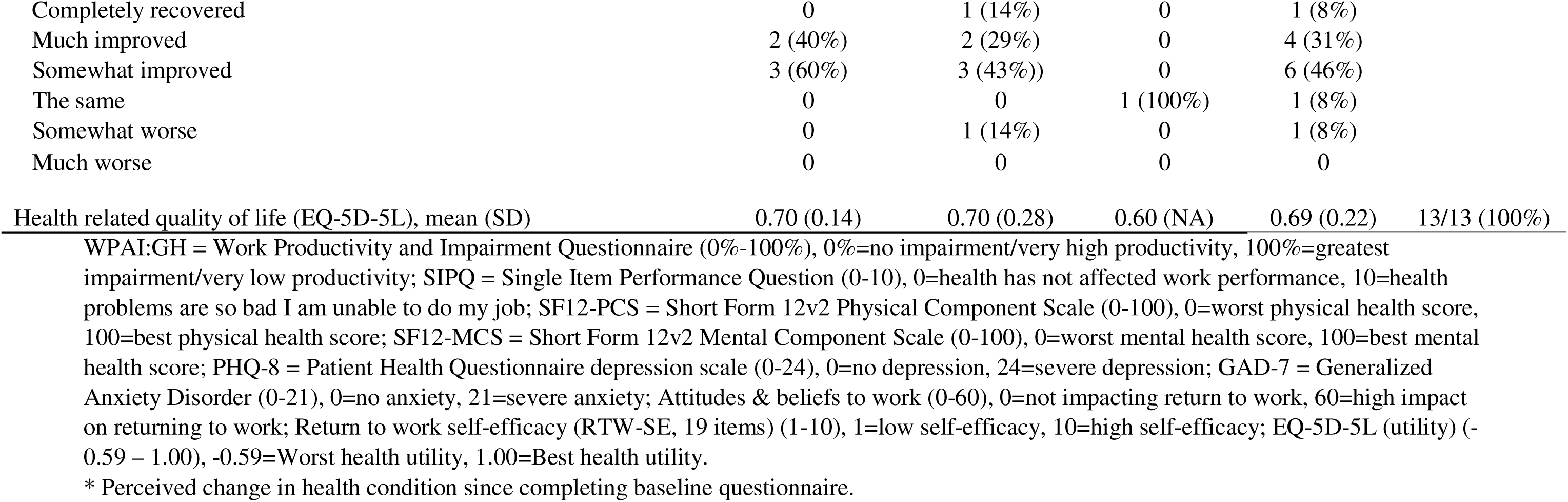
Summary of six-week follow-up data

Overall, SMS text message responsiveness was low, with 12/19 (63%) participants responding to the first message at Week 2 and 11/19 (58%) at Week 4. Subsequent messages had poor response rates, with only 42% (8/19) of participants providing a response to RTW (message 2 and 3) at Week 2 and 32% (6/19) at Week 4 (Table 4).

**Table 4:**
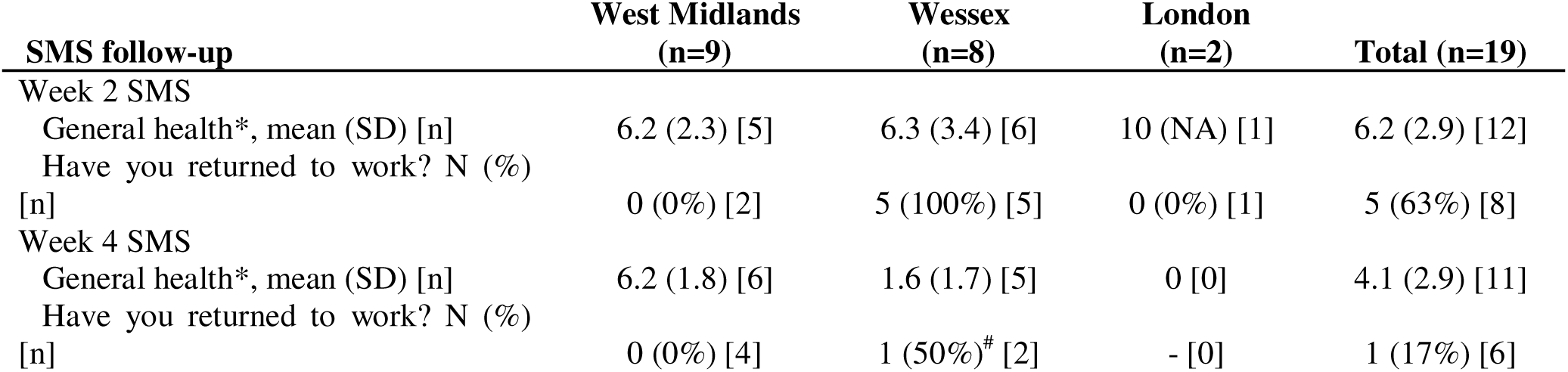
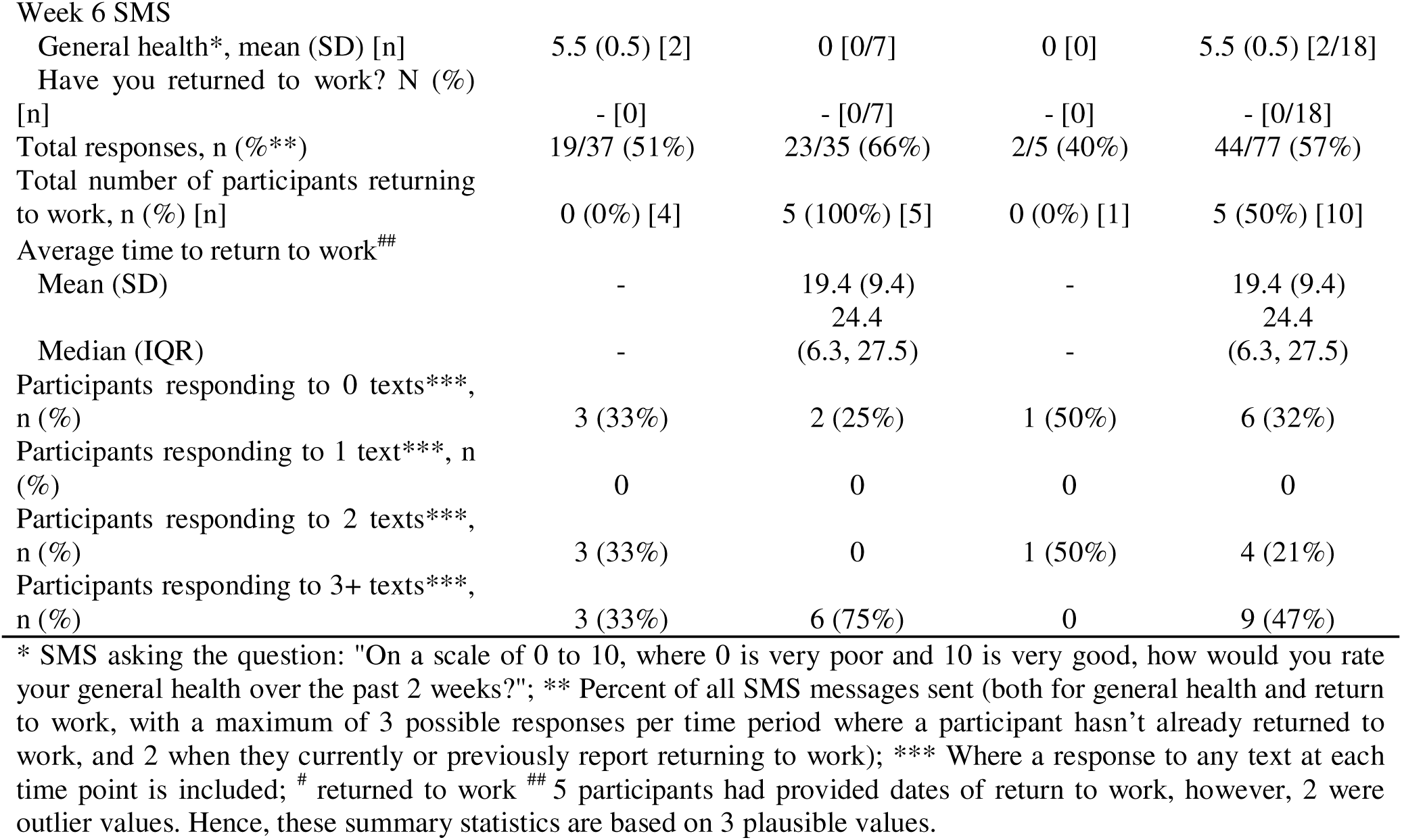
SMS Text messaging data

| **SMS follow-up** | **West Midlands**<br>**(n=9)** | **Wessex**<br>**(n=8)** | **London**<br>**(n=2)** | **Total (n=19)** |
| --- | --- | --- | --- | --- |
| Week 2 SMS |  |  |  |  |
| General health*, mean (SD) [n] | 6.2 (2.3) [5] | 6.3 (3.4) [6] | 10 (NA) [1] | 6.2 (2.9) [12] |
| Have you returned to work? N (%) |  |  |  |  |
| [n] | 0 (0%) [2] | 5 (100%) [5] | 0 (0%) [1] | 5 (63%) [8] |
| Week 4 SMS |  |  |  |  |
| General health*, mean (SD) [n] | 6.2 (1.8) [6] | 1.6 (1.7) [5] | 0 [0] | 4.1 (2.9) [11] |
| Have you returned to work? N (%) |  |  |  |  |
| [n] | 0 (0%) [4] | 1 (50%)<sup>#</sup> [2] | - [0] | 1 (17%) [6] |

**Table 4: SMS Text messaging data**
|  |  |  |  |  |
| --- | --- | --- | --- | --- |
| Week 6 SMS |  |  |  |  |
| General health*, mean (SD) [n] | 5.5 (0.5) [2] | 0 [0/7] | 0 [0] | 5.5 (0.5) [2/18] |
| Have you returned to work? N (%) |  |  |  |  |
| [n] | - [0] | - [0/7] | - [0] | - [0/18] |
| Total responses, n (%**) | 19/37 (51%) | 23/35 (66%) | 2/5 (40%) | 44/77 (57%) |
| Total number of participants returning to work, n (%) [n] | 0 (0%) [4] | 5 (100%) [5] | 0 (0%) [1] | 5 (50%) [10] |
| Average time to return to work<sup>##</sup> |  |  |  |  |
| Mean (SD) | - | 19.4 (9.4) | - | 19.4 (9.4) |
|  |  | 24.4 |  | 24.4 |
| Median (IQR) | - | (6.3, 27.5) | - | (6.3, 27.5) |
| Participants responding to 0 texts***, n (%) | 3 (33%) | 2 (25%) | 1 (50%) | 6 (32%) |
| Participants responding to 1 text***, n (%) | 0 | 0 | 0 | 0 |
| Participants responding to 2 texts***, n (%) | 3 (33%) | 0 | 1 (50%) | 4 (21%) |
| Participants responding to 3+ texts***, n (%) | 3 (33%) | 6 (75%) | 0 | 9 (47%) |

\* SMS asking the question: "On a scale of 0 to 10, where 0 is very poor and 10 is very good, how would you rate your general health over the past 2 weeks?"; \*\* Percent of all SMS messages sent (both for general health and return to work, with a maximum of 3 possible responses per time period where a participant hasn't already returned to work, and 2 when they currently or previously report returning to work); \*\*\* Where a response to any text at each time point is included; # returned to work <sup>##</sup> 5 participants had provided dates of return to work, however, 2 were outlier values. Hence, these summary statistics are based on 3 plausible values.

## Semi-structured interview findings

Analysis of data generated within the interviews explored participants’ experiences of being recruited to and participating in the WAVE feasibility study. Five participants were interviewed. Four participants were female and one male, aged from 45 to 70 years (mean age: 55), with variation in occupation, and reasons for receiving a fit note from their GP (Table 5). The length of each interview ranged between 30 minutes and 63 minutes (average: 48 minutes).

**Table 5:** Interview study participant characteristics

| Gender | Occupation type | Reason for fit note | No. of VSW consultations at time of interview |
| --- | --- | --- | --- |
| Female | Youth work | MSK | Three |
| Male | Emergency services | MSK | Did not consult with a VSW |
| Female | Healthcare work | MH | Two |
| Female | Emergency services | MH | Two |
| Female | Healthcare work | MH | Two |

MSK: Musculoskeletal conditions. MH: Mental health conditions

Three main themes were identified through the thematic analysis with full extracts reported in Supplementary Table 3:

1. Work absence and concerns about return to work
2. Views towards feasibility study processes
3. Acceptability and perceived value of the VSW service.

### Work absence and concerns about return to work

Participants reported concerns around work absence and RTW including workplace stress negatively impacting on health, adaptations to work routines as a result of COVID-19 adding additional pressure for some, and separation of work and home life being difficult leading to further negative health impacts. Support in the workplace was reported to be variable; some participants expressed concern about not receiving appropriate support upon returning whereas others reported accessing occupational health services at work and being well supported in their RTW planning.

*We’ve got a really good Occupational Health department. I’d actually self-referred to them before I went off because of the issues. I sort of self-referred in terms of ‘are there any things that I could or should be doing to make me deal with this better’, and as it turned out I think everyone including the line managers agree that it’s a situational thing. It’s not something about me. So yeah, I have seen occupational health, a counsellor from there. Gosh, probably about six or seven times over the last two or three months. (Female participant, in their 70s)*

A common concern was that the participants’ health condition would continue to prevent them fulfilling their job role and that they were unable to consider RTW planning until their health situation had improved. Despite these concerns, all participants expressed a desire to RTW noting the positive impact work had for them.

*Int: How important is it that you can actually return to work in the future to you?*

*P: Yes, for my sanity really. You know it’s a job I’ve done since I was sixteen. I only have eight years left before I can retire…and this is not the way I would have wanted to end my career really…it would be a sad way to leave. (Female participant, in their 50s)*

### Theme 2: Views towards feasibility study processes

All participants found it acceptable to be invited into the feasibility study. It was reported that first being alerted to the study by receiving a study pack through the post was acceptable, regardless of whether this was sent by their GP surgery or directly from the university. Some reported that they would not have felt it appropriate for their GP to explain the study to them in their consultation prior to receiving the study pack, as they would have had difficulty taking the information on board. Participants suggested that the WAVE study and intervention was not only acceptable in the context of the COVID-19 pandemic, but potentially of even greater relevance during this time, given the additional work pressures and shared concerns people were experiencing as a result of COVID.

*I think (WAVE) is acceptable, definitely, and because we’re in such a stressful time, albeit we seem to be coming out the other end now, but we don’t know where it’s going to go, but it’s good that you can do it when most people are under stress, not just certain people. Most people are under the same sort of stress. (Female participant, in their 50s)*

All participants also reported that the patient information and consent form received as part of the study pack were clear, and that based on this information they understood the aims and purpose of WAVE. The baseline questionnaire was reported to be acceptable in its use of language and length of the questionnaire. Whilst participants reported that, in general, they found it straightforward to complete, four participants reported either having difficulty in answering some questions with binary response options, or feeling that some of the questions were not applicable to them:

*Some of the questions, because I think a lot of it was about physical things, it didn’t sort of directly apply to me, anything like asking ‘do you do heavy lifting?’, and all that sort of thing. So for me I suppose it was a mental health thing more than a physical. A lot of it was easy to fill in because it wasn’t really kind of applicable in a way. (Female participant, in their 50s)*

In relation to the receipt of SMS text messages, four participants reported being uncertain at the time when the messages were received about who had sent them, due to the sender being displayed only as mobile phone number, without mention of the WAVE study. This uncertainty led to concerns that the text messages may have been sent from participants’ workplaces:

*I got sent a couple of texts and I thought ‘oh no this is from occupational health and I’m not going to answer it because I don’t know…’ as it was an unknown number and I thought ‘oh I don’t know this number, I’m not going to just answer these random questions’. (Female participant, in their 40s)*

### Theme 3: Acceptability and perceived value of the VSW service

One participant did not recall being contacted by a VSW. The other four participants reported being initially contacted by the VSW via telephone to arrange a suitable time for a consultation, following which all consultations were carried out via telephone. The participants reported finding this method of contact acceptable, particularly in light of social distancing guidance at the time due to COVID-19, and they felt able to effectively build rapport with the VSW via telephone. There was, however, the suggestion by a few of the participants that face-to-face contact could have further improved their experience, making it easier to ‘open up’.

Whilst only one participant who had consulted with the VSW had returned to work at the time of interview, they all reported feeling that the VSW service had been beneficial to them. Consulting with the VSW was seen to have value in several different ways; for instance, participants reported the benefits of interpersonal support provided by the VSW and having someone to talk to about their concerns. Continuity of support and ongoing contact provided by the VSW was also highlighted as valuable.

*I found it really useful for talking through some of the things that perhaps at the start of the call I hadn’t actually realised were playing on my mind that much, which was useful and we sort of talked through some things that I could do to try and ease my mind about that really. It is nice being able to talk to somebody about things. (Female participant, in their 70s)*

The support provided by VSWs in developing an action plan for RTW was another aspect of the intervention that was highlighted as valuable. However, the timing of RTW discussions was highlighted, with these conversations seen as being of lesser use early in the participant’s work absence, if they did not feel close to considering RTW. Whilst participants who received the VSW service all reported finding it beneficial, it was felt that support from a VSW would be of particular benefit to individuals who are not currently receiving support from their workplace, or do not have access to an occupational health department. Those who already had access to this support reported some overlaps between the VSW service and the support they were already receiving; for instance, where a phased return or workplace adjustments had already been discussed:

*She sent me an action plan through, and you know, she said ‘Well okay so if that’s what you’re worried about (the timeframe for returning to full duties), then how can you find that out?’; and she said ‘Could you ask your line manager?’ and I sort of said I could do, but I don’t really want to. I don’t know why, I just didn’t feel entirely comfortable…so what came out of it is that I said, ‘I need to understand really myself exactly what the policy says.’ So yeah, what we’d agreed is I either need to look that up myself or next time I see my counsellor, to go through that properly with her…so yeah, we’d talked through the various ways of how I could sort of put my mind at rest really. (Female participant, in their 40s)*

For participants who did not feel that their workplace had provided enough support to them, it was reported that support from the VSW could ‘add weight’ to discussions with work. The VSW had not directly contacted any of the participants’ workplaces, but some did report having discussed this, and all felt that this would be acceptable were it deemed necessary. However, some concerns were expressed about workplaces not being receptive to communication with the VSW, particularly in relation to issues of confidentiality and adherence to General Data Protection Regulations (GDPR).

## Discussion

The WAVE feasibility study explored the delivery of a VA intervention for adults in primary care certified absent from work for at least two weeks. The study experienced significant operational challenges, with the start of recruitment coinciding with the first wave of the COVID-19 vaccine roll-out by general practices and national lockdowns (December 2020 to March 2021). Despite this, the study met the two stop/go criteria, recruiting sufficient participants who were eligible and expressed an interest in the study (19/30 (58%)) and sufficient participants engaging with the VA intervention measured as having at least one contact with the VSW (16/19 (84%)). Questionnaire completion was good at both baseline and six-weeks’ follow-up. The use of SMS text messaging to gather shorter-term data on RTW was less successful, interviews with participants indicated that they were uncertain who had sent the messages as there was no mention of the WAVE study with the phone number. Analysis of the qualitative data indicated that the methods of inviting participants into the study were feasible and acceptable, and the study information was clearly understood. Telephone contact with the VSW was seen as acceptable and did not pose a barrier to building rapport, indicating that this would also be an appropriate form of contact for future delivery of a VA intervention. Furthermore, this type of intervention was seen as having even greater relevance given additional work pressures during the pandemic.

## Strengths and limitations of the study

This is the first study to test the feasibility of delivering a VA intervention to patients who present in primary care, regardless of their health condition. Previous research in this area has focused on specific health conditions (16,32,33). Focusing on one health condition may mean that those with other comorbid conditions are unable to access vocational advice. The WAVE intervention has the potential to address this gap in service provision given the intervention appears suitable for a wider primary care population. A further strength of the WAVE feasibility study is that it has tested three methods of participant recruitment. The subsequent learning from trialling these methods helped to identify the optimal recruitment strategy for a future trial i.e. one which focuses on automated identification and invitation as far as possible. Additionally, the mixed methods approach utilising the interview data reported here and recordings of intervention delivery reported elsewhere (14) is a key strength, since it has allowed the examination of not only the recruitment numbers from the different methods tested but also the fidelity of the delivery of the intervention and participants’ experiences of being invited to join the study and engage with the WAVE intervention. These findings can usefully inform the development of the methods for a future trial and also the WAVE intervention itself, to ensure that it meets the needs of participants in supporting them to RTW after a period of absence.

The study was limited by low recruitment (19 of the targeted 30 participants (63%)), and fewer participants agreed to take part in the interviews than was anticipated. This is set within the broader context of when the study was conducted. Our study was constrained by the unprecedented impacts of COVID-19 which influenced not only primary care, where the study was delivered, but also the wider context of work where there was a shift in working conditions for many people (34,35). The delivery of primary care moved to a predominantly online service, with changes made to many clinical and administrative processes including moving fit notes to an online request system when previously they required consultation with a clinician (11). Recruitment through consultations (Methods A and B) was significantly impacted by this change and recruitment methods were revised to allow identification of potential participants outside the consultation (Method C).

## Comparison with wider literature

COVID-19 has resulted in a prolonged period of exceptional pressure on primary care services which have moved to provide care in different ways, including remote consulting (36). Research such as the WAVE feasibility study, whilst impacted by this rapid shift, is still relevant and arguably more so as the recent report “Health is everyone’s business” highlights the need for improved vocational advice and support to be offered as part of the economic recovery plans (12). The report focuses on the need to support new occupational health models and make greater use of technology to support small and medium-sized enterprises to access occupational health services (12). The WAVE feasibility study responds to both these aspects: the VA intervention delivered in this feasibility study represents a novel model and the delivery of the service via telephone and videoconference fulfils the need for a novel approach to delivery, making good use of technology to support this.

There are continued concerns over the future availability of a responsive multi-disciplinary occupational health workforce (37). Government plans to respond to this perceived gap include considering methods to promote the expansion of clinical roles and improving occupational health multidisciplinary workforce models which capture both clinical and non-clinical roles and developing new training and career transition pathways (12). Conducting research such as the WAVE study supports the development of evidence-based models for provision of vocational advice and support for those without access to occupational health.

## Implications

The WAVE feasibility study highlights the challenges of working within changing clinical (primary care) and social environments (COVID-19). However, the findings align with the UK Government emphasis on improving vocational advice as part of economic recovery plans and as such further work is warranted in exploring how vocational advice can be successfully integrated into healthcare systems.

## Conclusions

The WAVE feasibility study demonstrated that delivering vocational advice in primary care is feasible, participants were successfully identified and recruited to the study and participants engaged in the VA intervention. Assessment of data collection processes indicated that response rates were acceptable, and data completion was very good. Importantly, participants reported that their experience of invitation and recruitment to the study was acceptable and that the VA intervention was useful in supporting them to RTW. The study has identified areas for refinement of recruitment strategies and indicated that clearer communication is required to maximise the potential impact. Progression to a full WAVE trial is indicated taking account of these findings and the implications noted.

## Declarations

### Ethical approval

Ethical approval was granted by National Research Ethics Service (NRES) Committee West of Scotland Research Ethics Committee (REC) 5, September 2020 (REC reference: 20/WS/0127).

### Availability of data and materials

All data are available on request, following Keele University’s data request process, by contacting the first author and.

### Author contributions

GW-J, IM, KW-B, CC, JP and NEF developed to concept of the WAVE study. NEF, GW-J, IM, KW-B, CC, JP GS, CC-G, SJ, ML, GM and SL obtained funding. GS, CC-G, IM, KWB, GW-J designed and developed the intervention and delivered training and mentoring to the vocational support workers employed. KB, ML, SJ, GM, BS, CC-G led and undertook all the analyses. KC, VP, CL and SL provided study management expertise and JP provided PPIE input.

## Data Availability

All data are available on request, following Keele Universitys data request process, by contacting the first author and.

## Acknowledgments

The team would like to thank the participants, the vocational support workers, the general practices and the Clinical Research Networks who all participated and supported in the delivery of this feasibility study. The team would also like to thank all those who contributed to the PPIE meetings which supported the development of the study from the initial development stages through to completion, in particular JP who led the PPIE throughout.

