## Supplementary information for "Can vocational advice be delivered in primary care? The Work And Vocational advicE (WAVE) mixed method single arm feasibility study (NCT04543097)"

**WAVE Feasibility study supplementary information**

**Supplementary Table 1: Full participant characteristics at baseline**

| **Baseline questionnaire** | **Staffordshire (n=9)** | **Wessex (n=8)** | **London (n=2)** | **Total (n=19)** | **Completeness** |
| --- | --- | --- | --- | --- | --- |
| Age, mean (SD) | 54.2 (15.4) | 54.6 (10.1) | 38.9 (10.4) | 52.8 (13.2) | 19/19 (100%) |
| Sex (Female), n (%) | 6 (67%) | 4 (50%) | 1 (50%) | 11 (58%) | 19/19 (100%) |
| Level of education, n (%) |  |  |  |  | 19/19 (100%) |
| O Levels/GCSEs | 6 (67%) | 7 (88%) | 0 (0%) | 13 (68%) |  |
| A Levels | 2 (22%) | 3 (38%) | 1 (50%) | 6 (32%) |  |
| Vocational training certificates | 5 (56%) | 8 (100%) | 1 (50%) | 14 (74%) |  |
| Higher professional qualifications | 4 (44%) | 2 (25%) | 1 (50%) | 7 (37%) |  |
| Main health condition causing time off work, n (%) |  |  |  |  | 16/19 (84%) |
| Aches and pains | 2 (29%) | 2 (25%) | 1 (100%) | 5 (31%) |  |
| Mental health problems | 5 (71%) | 2 (25%) | 0 | 7 (44%) |  |
| Other health condition | 0 | 4 (50%) | 0 | 4 (25%) |  |
| Number of days absence, mean (SD); median [IQR] | 14.9 (14.6); 15 [0.8, 23.5] | 18.4 (12.6); 16 [9.5, 26.3] | 97 (117.4); 97 [55.5, 138.5] | 25.6 (40.5);  16 [8.5, 25] | 18/19 (95%) |
| Work interference [WPAI:GH], mean % (SD) |  |  |  |  |  |
| Absenteeism | 100 (0) | 91.4 (22.7) | 100 (NA) | 96.3 (15) | 16/19 (84%) |
| Presenteeism | 100 (0) | 90 (NA) | 90 (NA) | 95 (5.8) | 4/19 (21%) |
| Work productivity | 100 (0) | 94 (NA) | NA | 98 (3.5) | 3/19 (16%) |
| Activity impairment | 63.3 (30) | 71.3 (23.6) | 50 (14.1) | 65.3 (25.9) | 19/19 (100%) |
| Work performance [SIPQ], mean (SD) | 5.8 (3.8) | 3.9 (3.6) | 6.0 (4.2) | 5.0 (3.7) | 19/19 (100%) |
| SF12-PCS, mean (SD) | 36.0 (13.0) | 38.0 (15.8) | 33.2 (10.8) | 36.6 (13.5) | 18/19 (95%) |
| SF12-MCS, mean (SD) | 31.1 (10.1) | 35.8 (7.6) | 40.2 (3.2) | 34.2 (8.8) | 18/19 (95%) |
| Depression (PHQ-8), mean (SD) | 14.0 (5.3) | 8.9 (6.2) | 8.0 (7.1) | 11.2 (6.1) | 19/19 (100%) |
| Anxiety (GAD-7), mean (SD) | 13.1 (4.3) | 8.1 (6.4) | 9.5 (6.4) | 10.6 (5.7) | 19/19 (100%) |
| Attitudes & beliefs to work, mean (SD) | 40.1 (4.3) | 37.0 (4.2) | 42.0 (5.7) | 39.0 (4.5) | 19/19 (100%) |
| Return to work self-efficacy (RTW-SE), mean (SD) | 3.9 (2.2) | 4.8 (2.3) | 6.5 (0.9) | 4.5 (2.2) | 19/19 (100%) |
| Physical activity level (GPPAQ3), n (%) |  |  |  |  | 17/19 (89%) |
| Inactive | 3 (33%) | 1 (17%) | 0 | 4 (24%) |  |
| Moderately inactive | 2 (22%) | 2 (33%) | 1 (50%) | 5 (29%) |  |
| Moderately active | 2 (22%) | 1 (17%) | 0 | 3 (18%) |  |
| Active | 2 (22%) | 2 (33%) | 1 (50%) | 5 (29%) |  |
| Use and content of other services providing vocational advice, n (%) |  |  |  |  |  |
| Vocational advisor | 0 | 1 (17%) | 0 | 1 (8%) | 12/19 (63%) |
| GP | 9 (100%) | 5 (71%) | 2 (100%) | 16 (89%) | 18/19 (95%) |
| My organisation's occ. Health department | 5 (71%) | 3 (43%) | 1 (100%) | 9 (60%) | 15/19 (79%) |
| Practice nurse | 1 (14%) | 1 (17%) | 0 | 2 (15%) | 13/19 (68%) |
| Physiotherapist | 1 (17%) | 3 (50%) | 2 (100%) | 6 (43%) | 14/19 (74%) |
| Other | 3 (60%) | 3 (60%) | 0 | 6 (60%) | 10/19 (53%) |
| Working hours, n (%) |  |  |  |  | 18/19 (95%) |
| Full time (≥35 hours per week) | 5 (63%) | 6 (75%) | 2 (100%) | 13 (72%) |  |
| Part time (<35 hours per week) | 3 (38%) | 2 (25%) | 0 | 5 (28%) |  |
| Work characteristics, n (%) |  |  |  |  |  |
| Kneeling/squatting for >1 hour per day | 2 (22%) | 2 (25%) | 1 (50%) | 5 (26%) | 19/19 (100%) |
| Climbing a ladder | 1 (11%) | 1 (13%) | 1 (50%) | 3 (16%) | 19/19 (100%) |
| Climbing up/down stairs >30 times per day | 2 (22%) | 3 (38%) | 1 (50%) | 6 (32%) | 19/19 (100%) |
| Digging or shovelling | 1 (11%) | 1 (13%) | 1 (50%) | 3 (16%) | 19/19 (100%) |
| Lifting weights of ≥10kg by hand | 2 (22%) | 2 (25%) | 1 (50%) | 5 (26%) | 19/19 (100%) |
| Standing/walking for most of the day | 3 (33%) | 2 (25%) | 2 (100%) | 7 (37%) | 19/19 (100%) |
| Standing/walking for >3 hours at a time | 3 (33%) | 2 (25%) | 2 (100%) | 7 (37%) | 19/19 (100%) |
| Hard physical work that makes you hot/sweaty | 1 (11%) | 3 (38%) | 1 (50%) | 5 (26%) | 19/19 (100%) |
| What is the gross total income from all sources per week? n (%) |  |  |  |  | 19/19 (100%) |
| £0-£99 (£0-£5,199 per year) | 0 | 0 | 0 | 0 |  |
| £100-£149 (£5,200-£7,799 per year) | 0 | 0 | 0 | 0 |  |
| £150-£249 (£7,800-£12,999 per year) | 0 | 0 | 0 | 0 |  |
| £250-£349 (£13,000-£18,199 per year) | 0 | 0 | 0 | 0 |  |
| £350-£449 (£18,200-£23,399 per year) | 1 (11%) | 1 (13%) | 0 | 2 (11%) |  |
| £450-£599 (£23,400-£31,199 per year) | 1 (11%) | 0 | 0 | 1 (5%) |  |
| £500-£749 (£31,200-£38,999 per year) | 2 (22%) | 0 | 1 (50%) |  |  |
| £750 or more (£39,00 or more per year) | 5 (56%) | 4 (50%) | 1 (50%) | 10 (53%) |  |
| Prefer not to say | 0 | 3 (38%) | 0 | 3 (16%) |  |
| Satisfaction with work, mean (SD) | 5.2 (3.0) | 6.1 (2.3) | 4 (2.8) | 5.5 (2.7) | 19/19 (100%) |
| How do you commute to work? n (%) |  |  |  |  | 18/19 (95%) |
| Walk | 0 | 0 | 0 | 0 |  |
| Cycle | 0 | 0 | 0 | 0 |  |
| Public transport | 0 | 0 | 1 (100%) | 1 (6%) |  |
| Car (private or shared) | 8 (89%) | 6 (75%) | 0 | 14 (78%) |  |
| N/A (e.g. work from home) | 1 (11%) | 2 (25%) | 0 | 3 (17%) |  |
| How long is your commute to work (minutes)? |  |  |  |  | 19/19 (100%) |
| Mean (SD) [median] | 28.1 (37.3) [15] | 27.2 (11.8)  [22.5] | 40 (28.3)  [40] | 29.3 (27.7) [20] |  |
| Not applicable | 1 (11%) | 2 (25%) | 0 | 3 (16%) |  |
| Health related quality of life (EQ-5D-5L), mean (SD) | 0.56 (0.2) | 0.56 (0.3) | 0.63 (0.05) | 0.57 (0.24) | 19/19 (100%) |
| IQR: Intuartile range [25%, 75%]. WPAI:GH = Work Productivity and Impairment Questionnaire (0%-100%), 0%=no impairment/very high productivity, 100%=greatest impairment/very low productivity; SIPQ = Single Item Performance Question (0-10), 0=health has not affected work performance, 10=health problems are so bad I am unable to do my job; SF12-PCS = Short Form 12v2 Physical Component Scale (0-100), 0=worst physical health score, 100=best physical health score; SF12-MCS = Short Form 12v2 Mental Component Scale (0-100), 0=worst mental health score, 100=best mental health score; PHQ-8 = Patient Health Questionnaire depression scale (0-24), 0=no depression, 24=severe depression; GAD-7 = Generalized Anxiety Disorder (0-21), 0=no anxiety, 21=severe anxiety; Attitudes & beliefs to work (0-60), 0=not impacting return to work, 60=high impact on returning to work; Return to work self-efficacy (RTW-SE, 19 items) (1-10); 1=low self-efficacy, 10=high self-efficacy; EQ-5D-5L (utility) (-0.59 – 1.00), -0.59=Worst health utility, 1.00=Best health utility. | | | | |  |

**Supplementary Table 2: Full participant characteristics at six weeks**

|  | **Staffordshire (n=5)** | **Wessex (n=7)** | **London (n=1)** | **Total (n=13)** | **Completeness** |
| --- | --- | --- | --- | --- | --- |
| Number of days absence, mean (SD) ; median [IQR] | 22.0 (11.0) 30 [10, 30] | 20.1 (21.1)  15 [0, 42] | 0 | 19.3 (17.3)  15 [0, 30] | 13/13 (100%) |
| Current work status, n (%) |  |  |  |  | 13/13 (100%) |
| Doing your usual job | 1 (20%) | 4 (57%) | 1 (100%) | 6 (46%) |  |
| On paid/annual leave | 2 (40%) | 0 | 0 | 2 (15%) |  |
| Working fewer hours | 0 | 1 (14%) | 0 | 1 (8%) |  |
| Doing lighter duties | 0 | 0 | 0 | 0 |  |
| On paid sick leave | 2 (40%) | 2 (29%) | 0 | 4 (31%) |  |
| On unpaid sick leave | 0 | 0 | 0 | 0 |  |
| Work interference [WPAI:GH], mean % (SD) |  |  |  |  |  |
| Absenteeism | 78.1 (37.9) | 28.6 (48.8) | 0 (NA) | 39.5 (49) | 11/13 (85%) |
| Presenteeism | 40 (14.1) | 52 (23.9) | 50 (NA) | 48.8 (19.6) | 8/13 (62%) |
| Work productivity | 83.6 (23) | 52 (23.9) | 50 (NA) | 59.6 (24.9) | 8/13 (62%) |
| Activity impairment | 42 (22.8) | 52.9 (33.5) | 50 (NA) | 48.5 (27.6) | 13/13 (100%) |
| Work performance [SIPQ], mean (SD) | 9.2 (1.8) | 6.4 (3.4) | 5 (NA) | 7.4 (3.0) | 13/13 (100%) |
| SF12-PCS, mean (SD) | 42.7 (11.8) | 38.4 (15.9) | 41.8 (NA) | 40.3 (13.3) | 13/13 (100%) |
| SF12-MCS, mean (SD) | 34.8 (10.9) | 43.0 (12.0) | 38.2 (NA) | 39.5(11.3) | 13/13/(100%) |
| Depression [PHQ-8], mean (SD) | 9.8 (6.4) | 8.0 (6.7) | 11 (NA) | 8.9 (6.1) | 13/13 (100%) |
| Anxiety [GAD-7], mean (SD) | 8.0 (5.2) | 4.3 (3.5) | 14 (NA) | 6.5 (4.8) | 13/13 (100%) |
| Attitudes & beliefs to work, mean (SD) | 38 (7.2) | 36.9 (3.5) | 43 (NA) | 37.8 (5.1) | 13/13 (100%) |
| Return to work self-efficacy [RTW-SE], mean (SD) | 5.6 (2.1) | 5.6 (2.5) | 7 (NA) | 5.7 (2.2) | 13/13 (100%) |
| Physical activity level [GPPAQ3], n (%) |  |  |  |  | 12/13 (92%) |
| Inactive | 0 | 2 (29%) | 0 | 2 (17%) |  |
| Moderately inactive | 1 (25%) | 2 (29%) | 0 | 3 (25%) |  |
| Moderately active | 0 | 1 (14%) | 0 | 1 (8%) |  |
| Active | 3 (75%) | 2 (29%) | 1 (100%) | 6 (50%) |  |
| Use and content of other services providing vocational advice, n (%) |  |  |  |  |  |
| Vocational advisor | 1 (20%) | 2 (29%) | 1 (100%) | 4 (31%) | 13/13 (100%) |
| GP | 5 (100%) | 4 (57%) | 1 (100%) | 10 (77%) | 13/13 (100%) |
| My organisation's occ. Health department | 4 (80%) | 1 (14%) | 1 (100%) | 6 (46%) | 13/13 (100%) |
| Practice nurse | 0 | 2 (29%) | 1 (100%) | 3 (23%) | 13/13 (100%) |
| Physiotherapist | 0 | 2 (29%) | 0 | 2 (15%) | 13/13 (100%) |
| Other | 1 (20%) | 3 (50%) | 0 | 4 (40%) | 10/13 (77%) |
| Working hours, n (%) |  |  |  |  | 11/13 (85%) |
| Full time (≥35 hours per week) | 2 (67%) | 5 (71%) | 1 (100%) | 8 (73%) |  |
| Part time (<35 hours per week) | 1 (33%) | 2 (29%) | 0 | 3 (27%) |  |
| Work characteristics, n (%) |  |  |  |  |  |
| Kneeling/squatting for >1 hour per day | 0 | 2 (29%) | 0 | 2 (17%) | 12/13 (92%) |
| Climbing a ladder | 0 | 1 (14%) | 0 | 1 (8%) | 12/13 (92%) |
| Climbing up/down stairs >30 times per day | 0 | 1 (14%) | 0 | 1 (8%) | 12/13 (92%) |
| Digging or shovelling | 0 | 1 (14%) | 0 | 1 (8%) | 12/13 (92%) |
| Lifting weights of ≥10kg by hand | 1 (25%) | 2 (29%) | 0 | 3 (25%) | 12/13 (92%) |
| Standing/walking for most of the day | 2 (50%) | 3 (43%) | 1 (100%) | 6 (50%) | 12/13 (92%) |
| Standing/walking for >3 hours at a time | 0 | 2 (29%) | 1 (100%) | 3 (25%) | 12/13 (92%) |
| Hard physical work that makes you hot/sweaty | 1 (25%) | 2 (29%) | 0 | 3 (25%) | 12/13 (92%) |
| Satisfaction with work, mean (SD) | 6.0 (1.4) | 6.0 (1.8) | 8 (NA) | 6.2 (1.6) | 12/13 (92%) |
| Perceived change in health condition*, n (%) |  |  |  |  | 13/13 (100%) |
| Completely recovered | 0 | 1 (14%) | 0 | 1 (8%) |  |
| Much improved | 2 (40%) | 2 (29%) | 0 | 4 (31%) |  |
| Somewhat improved | 3 (60%) | 3 (43%)) | 0 | 6 (46%) |  |
| The same | 0 | 0 | 1 (100%) | 1 (8%) |  |
| Somewhat worse | 0 | 1 (14%) | 0 | 1 (8%) |  |
| Much worse | 0 | 0 | 0 | 0 |  |
| Health related quality of life (EQ-5D-5L), mean (SD) | 0.70 (0.14) | 0.70 (0.28) | 0.60 (NA) | 0.69 (0.22) | 13/13 (100%) |

**Supplementary Table 3: Extracts from interviews with participants**

| **Theme 1: Work absence and concerns about RTW** | **Theme 2: Views towards feasibility trial processes** | **Theme 3: Acceptability and perceived value of the VA intervention** |
| --- | --- | --- |
| *I’ve been finding work was becoming sort of increasingly stressful as time went on…the volume of work and the intensity really, that I found that I was literally just constantly on the go. You know, I’d generally work an eight or nine hour shift if I finished on time and throughout that entire eight or nine hours it would be literally just non-stop with one thing or another. So I’d be getting symptoms, like ridiculously tired, headaches, often getting very hot and cold and feeling like I’d got a temperature. You know, sort of general rundown sort of feeling…and I knew what it was, I felt that it was related to the stress and anxiety and being rundown. (Female participant in their 40s)* | *If the GP had mentioned it at the time of my consultation. I don’t think that would’ve been the right time because it’s a situation where you’re quite emotional and you’re talking about a lot of stuff and it’s important, isn’t it? So I don’t think I would really have taken it in that well or certainly probably not have understood what it was about. So yeah, I wouldn’t really have wanted the GP to mention it at the time I don’t think. (Female participant in their 40s)* | *Int: How did you find that that consultation worked over the phone?*  *P: Oh very well, I was very happy to just do a phone call, yes.*  *Int: Okay so in terms of communication and building a relationship, did you feel there were any barriers at all in doing it that way?*  *P: No, he was immediately really easy to talk to and he was really good.*  *(Female participant in their 50s)* |
| *Covid alone had a huge impact, a massive impact. Not the disease, the management, trying to manage for it had a massive impact. Then the Covid disease itself had a massive impact, then on top of that so many injuries because people were working trying to cover people off sick, so the degradation amongst the workforce was immense. We had more people, not necessarily off sick, but getting injured. And then as a result they had more people having to cover, hence why I’ve done more shifts.*  *(Male participant in their 40s)* | *I think (WAVE) is acceptable, definitely, and because we’re in such a stressful time, albeit we seem to be coming out the other end now, but we don’t know where it’s going to go, but it’s good that you can do it when most people are under stress, not just certain people. Most people are under the same sort of stress.*  *(Female participant in their 50s)* | *I think it’s hard for the person who’s being supported, it can be hard for them to be open when you’re talking over the phone… I believe that it would’ve been a better experience if I’d been able to do those sessions face to face.*  *(Female participant in their 40s)* |
| *Covid and working from home…we had huge IT problems so we couldn’t log on because everybody else was logging on so you'd be told try after 10 o’clock, try after 11, try after 12 and we weren’t getting any work done. You're also working with young people, so it was just by phone, it was also issues about learning new technology like Zoom, Facetiming, all those kind of things. And I just found from then, I was just so overwhelmed, with being allocated more and more cases…I was set up in the kitchen, so I saw it all the time, I'm in the kitchen, I can see my desk when I’m out in the back where we eat; and if I watch telly constantly walk past adding to my to-do list; I've forgotten this, I've forgotten that, or just carrying on working.*  *(Female participant in their 50s)* | *Yes, it was clear, yes about supporting me to get back to work, that was fine… I think they were pitched at the right level (Female participant in their 50s)* | *I found it really useful for talking through some of the things that perhaps at the start of the call I hadn’t actually realised were playing on my mind that much, which was useful and we sort of talked through some things that I could do to try and ease my mind about that really. It is nice being able to talk to somebody about things.*  *(Female participant in their 70s)* |
| *P: We discussed light duties, but my line manager just laughed and said, ‘We are where we are.’ It only takes one person off sick and we’re in trouble.*  *Int: Did you have any concerns about going back?*  *P: Completely, but what can you do?*  *Int: Is there any form of support available for you?*  *P: Ibuprofen and gaffer tape (laughs). There is no support because what do I do? Do I phone up? No one can change the situation, no one can change the job we do and I can’t go to work in a plaster cast*  *(Male participant in their 40s)* | *Some of the questions, because I think a lot of it was about physical things, it didn’t sort of directly apply to me, anything like asking ‘do you do heavy lifting?’, and all that sort of thing. So for me I suppose it was a mental health thing more than a physical. A lot of it was easy to fill in because it wasn’t really kind of applicable in a way.*  *(Female participant in their 50s)* | *I think it’s just keeping in touch, maintaining that contact really…he was really good, he was really supportive, and actually when I was thinking ‘oh I must go back’, he said ‘would you like me to call you before you go back?’ and I said ‘yes that would be really good if you could just call me the day before I go back to work’, because I was feeling really very apprehensive.*  *(Female participant in their 50s)* |
| *We’ve got a really good Occupational Health department. I’d actually self-referred to them before I went off because of the issues. I sort of self-referred in terms of ‘are there any things that I could or should be doing to make me deal with this better’, and as it turned out I think everyone including the line managers agree that it’s a situational thing. It’s not something about me. So yeah, I have seen occupational health, a counsellor from there. Gosh, probably about six or seven times over the last two or three months. I’m seeing her again this afternoon actually. (Female participant in their 70s)* | *I only had a couple (of text messages) but that wasn’t clear really. I haven't really replied to them because I didn’t feel they were relevant… I'm just trying to look for them. Oh I received three which was the XX of February, XX of February and XX of March. But that’s not clear who it’s from and in the scheme of things of getting anything, emails or you know, it’s just not clear enough, it’s just from a like a mobile number.*  *(Female participant in their 70s)* | *She sent me an action plan through, and you know, she said ‘Well okay so if that’s what you’re worried about (the timeframe for returning to full duties), then how can you find that out?’; and she said ‘Could you ask your line manager?’ and I sort of said I could do, but I don’t really want to. I don’t know why, I just didn’t feel entirely comfortable…so what came out of it is that I said, ‘I need to understand really myself exactly what the policy says.’ So yeah, what we’d agreed is I either need to look that up myself or next time I see my counsellor, to go through that properly with her…so yeah, we’d talked through the various ways of how I could sort of put my mind at rest really.*  *(Female participant in their 40s)* |
| *I mean the prospect of returning to work, especially for me, my job is a thinking about things job and obviously the reason I’m off is mental health related. So the thought of going back, it’s a bit like if someone had got a job where they’d broken their arm, but the job was a very physical job that involved putting boxes on high shelves or something. You know, you’re sort of worried that the part of you that needs to be working well to do that job isn’t working properly. So going back is quite scary and just figuring out what a phased or a structured return is like. (Female participant in their 50s)* | *I got sent a couple of texts and I thought ‘oh no this is from occupational health and I’m not going to answer it because I don’t know…’ as it was an unknown number and I thought ‘oh I don’t know this number, I’m not going to just answer these random questions’. (Female participant in their 40s)* | *The last time I spoke to her was two weeks ago and gosh, it was probably a month before I was due to return to work then. So it was not really at a stage where you could really start to plan details of a phased return plan. Because at that time I didn’t even know when I would be going back and at that time I remember saying to (the VSW) ‘If you said to me now do you think you’ll be ready to go back to work in three weeks’ time?’ I said ‘No, definitely not. Unless there’s a miracle in the meantime, I don’t think I’m going to be ready.’ So it was all very open-ended at the time, so possibly a little bit early at that time to be discussing those sorts of things too much.*  *(Female participant in their40s)* |
| *Int: You mentioned you went to the doctor the week after Christmas, was that when you were signed off for the last time, were you given the fit note at that point?*  *P: Yes, I haven't returned since. And that’s why I feel stuck because I don’t foresee being able to (RTW) because the op hasn’t gone ahead, I'm not on the medication to manage the arthritis.*  *Int: So at the minute you can’t really see an end to where you might be able to return?*  *P: No, and I’m trying to be positive about that it’s just…yes.*  *(Female participant in their 40s)* | *The texts were fine, although I wasn’t really quite sure whether I understood the question right, because the question was ‘how would you describe your general health?’, I think. And when I see general health I read that as saying sort of more physical health. If the question had been ‘How would you describe your mental health?’ I would’ve expected to say mental health. So I wasn’t really quite sure. So that was maybe a little bit ambiguous...I sort of answered it in terms of my general health and in actual fact since I’ve been off my general health has probably improved. Whereas if I’d sort of thought, ‘Right this is specifically in relation to my mental health and how I’m feeling’ then it would’ve been quite a different answer.*  *(Female participant in their 70s)* | *I do think it added but I think it would be even more beneficial to people who just didn’t have that at all. I can just think for some people, maybe working in retail or something where you just don’t have the network that we have…that would be so valuable.*  *(Female participant in their 70s)* |
| *Int: How important is it that you can actually return to work in the future to you?*  *P: Yes, for my sanity really. You know it’s a job I've done since I was sixteen. I only have eight years left before I can retire…and this is not the way I would have wanted to end my career really…it would be a sad way to leave.*  *(Female participant in their 50s)* |  | *I think sometimes (work managers) don’t believe or accept everything that’s in an occupational health assessment. And they don’t act quick enough on it, that’s been my experience. So with this (the VSW service), I think the support from this, you know would add weight to that…I still need that backing I think to make sure that they do it. (Female participant in their 70s)* |
|  |  | *if I’d had a manager that wasn’t helpful and just expected you to come back and start off where you left off, he said ‘I can get involved with speaking to your manager and talking about phased return’. So I could see he’d be very proactive if I wasn’t in such a fortunate situation.* |
|  |  | *I’m not sure how our work would take that, because obviously they’re quite protective and GDPR etcetera, I know they’ve had issues around that before...I think our work are very, very protective around that, so I don’t know how that would work for an outsider body contacting them unless they are pre-authorised.* |
